# The effects of maternal mental health in pregnancy on neonatal adversity: role of physical health and familial factors

**DOI:** 10.1101/2022.11.02.22281841

**Authors:** Emma Lin, Elah Wilson, Arad Kodesh, Stephen Z. Levine, Nathan Fox, Abraham Reichenberg, Vahe Khachadourian, Magdalena Janecka

## Abstract

**Background:** There exists substantial evidence of the effects of maternal mental health on birth outcomes. However, the roles of (1) comorbidity between mental and physical health, and (2) familial confounding underlying this association, remain unclear.

**Methods:** This cohort study included a random sample of 19.5% children born between January 1, 1997, through December 31, 2008, within a health maintenance organization (HMO) in Israel, as well as their parents and siblings born within the cohort years. Outcomes were ICD-9 diagnoses of neonatal adversity (birth complications and congenital anomalies), and exposure was parental mental health diagnosis – all ascertained through the HMO records. Odds ratios (ORs) and their 95% CIs for the associations between the maternal mental health diagnosis and neonatal adversity were calculated using logistic regression, iteratively adjusting for covariates (maternal age, child’s year of birth, socioeconomic status, number of physical health diagnoses). We also examined potential familial confounding using a negative control approach based on paternal exposure.

**Results:** In our sample of 74,533 children, 6,674 (9.1%) were born after birth complications and 14,569 (19.9%) with a congenital anomaly. Maternal mental health diagnosis around pregnancy was significantly associated with these measures of neonatal adversity, adjusting for demographic and socioeconomic factors (*birth complications*: OR=1.3 (1.2-1.4); p<0.001; *congenital anomalies:* OR=1.2 (1.1-1.3); p<0.001). These associations became attenuated and non-significant after further adjustment for the number of physical health diagnoses. In a joint model, maternal and paternal diagnosis of mental health disorder were independently associated with measures of neonatal adversity (*birth complications*: OR_mat_=1.3 (1.1-1.4); p < 0.001; OR_pat_=1.2 (1.1-1.3); p=0.004; *congenital anomalies*: OR_mat_=1.2 (1.1-1.3); p<0.001; OR_pat_=1.1 (1.0-1.2); p=0.01).

**Conclusions:** Physical health comorbidities and familial factors play a role in the associations between maternal mental health and neonatal adversity.

## Introduction

A diagnosis of maternal mental health disorder during pregnancy has been associated with several adverse birth outcomes in offspring, including prematurity^1^, stillbirth^2^ and congenital anomalies^2,3^. Unlike longer-term adversity, for example behavioral problems in offspring^4^, associations with birth outcomes are less likely to arise due to the psychosocial factors correlated with growing up with a parent with a mental health disorder. Consequently, several biological mechanisms have been proposed to account for the observational associations between maternal mental health and birth outcomes, predominantly focusing on the role of stress hormones, inflammatory factors or changes in placental fuction^5–7^. Nevertheless, the empirical evidence for the role of these factors remain limited, and how such factors could mediate the impact on child’s birth outcomes is not fully understood, leaving a possibility of background confounding by other maternal and/or other familial factors.

While many studies investigating the birth sequelae of maternal diagnosis of mental disorder in pregnancy implemented careful adjustment to control for a number of potential confounders, *e*.*g*. age at conception, socioeconomic status or substance use, the effects of many maternal factors, including measures of her physical health, remain unknown. This is despite substantial evidence for a pervasive comorbidity between mental and physical disorders^8^, including in pregnant women^9^ - which entails that pregnant women with diagnosed mental health conditions are more likely to present with a number of additional health problems. As many of these physical health disorders are themselves associated with neonatal adversity^10,11^, the extent to which they could contribute to the association between maternal mental illness and offspring outcomes remains unclear.

In addition, family-level characteristics associated with maternal mental illness, *e*.*g*. environmental pollutants, socioeconomic status, or their genetic underpinnings, could all introduce spurious association between maternal mental illness and neonatal adversity. One of the ways to account for such familial factors, even in the absence of detailed data on these potential confounders, is the use of a negative control of paternal exposure^12^, *i*.*e*. leveraging the fact that paternal exposures during pregnancy cannot directly influence the fetus. For example, only a potential increase in maternal, but not paternal levels of inflammation or stress hormones due to mental health issues can have direct negative health effects on the fetus. Therefore, observing an equal association between neonatal adversity and mental illness in either of the parents provides indirect evidence for the role of family-level factors, rather than direct influences of maternal mental illness on the fetus.

Better understanding of the mechanisms underlying the effects of maternal mental health in pregnancy on offspring outcomes is critical to identify potential targets for preventing neonatal and child adversity^13^. While maternal mental health in pregnancy requires attention irrespective of its putative effects on offspring, elucidating the role of background confounders in the observational associations may yield further modifiable factors that warrant attention. Therefore, we investigated the contribution of (a) maternal physical health, and (b) familial factors, to the observational associations between maternal mental health diagnosis and neonatal adversity, using a large, representative sample from Israel with detailed medical and demographic data.

## Methods

This study was approved by the institutional review board of the University of Haifa and the Helsinki Ethics Committee. Those bodies waived the need for informed consent because the study data were fully deidentified.

### Sample

We used a population-based cohort from Meuhedet, a large health maintenance organization (HMO) in Israel. We obtained data on a random sample of 19.5% (95,978) of all children born in the HMO between January 1, 1997, and December 31, 2008, as well as their siblings and parents. In order to ensure reliable coverage of parental health records before the child’s birth, we excluded all individuals born before January 1, 1999. Other exclusion criteria included: lack of at least one medical diagnosis in parental records (indicating administrative lapse in recording); maternal age at child’s birth lower than 13, or higher than 55, or paternal age lower than 13; non-singleton pregnancies; lack of covariate data. All children were followed up through January 2015.

### Exposures

Exposure was presence of *any* ICD-9 mental health diagnosis (ICD-9: 290-319) recorded within 635 days before child’s birth (spanning the entire pregnancy period and the preceding 12 months, assuming a term pregnancy), in the mother – period subsequently referred to as “around pregnancy”. Extending the exposure period to 12 month before pregnancy allowed us to ascertain chronic disorders for which the mother may not seek help during pregnancy itself, but which may still affect the fetus (*NB*. while this approach increases a probability of introducing/increasing familial confounding due to potential ascertainment of diagnoses that do not affect the fetus directly, this issue is addressed through paternal comparisons). In additional analyses we assessed the differential effects across prevalent mental health diagnoses in our sample^9^, *i*.*e*. anxiety, dissociative, and somatoform disorders (ICD-9: 300); personality disorders (ICD-9: 301); other disorders with specific symptoms (ICD-9: 307); depressive disorders (ICD-9: 311). All exposures were coded as binary variables, indicating presence/absence of the relevant ICD-9 codes in maternal health records. The same criteria were used to ascertain paternal diagnoses of mental health disorders.

### Outcome

The outcomes were two categories of neonatal adversity: *birth complications* (ICD-9: 660-669; 678-679, 760-779) and *congenital anomalies* of the child (ICD-9: 740-759). Both outcomes were coded as binary variables, indicating presence/absence of the relevant code in the child’s health records obtained from the HMO. We only considered diagnoses made within three years of child’s birth, allowing for the delay in the diagnosis of certain congenital anomalies, while ensuring no child could be administratively censored (end of the follow up) before age of three. In additional sensitivity analyses we only considered the ICD-9 outcome codes with a relatively low prevalence in the sample (<1.5%) to focus on the diagnoses more likely associated with significant adversity.

### Covariates

The covariates included: child’s year of birth, maternal age at child’s birth, family socioeconomic status (SES), and the number of distinct maternal medical comorbidities during the exposure period. Adjusting for the calendar year of child’s birth allowed us to mitigate against confounding due to temporal changes in the diagnostic rates of psychiatric disorders and neonatal adversity. Controlling for maternal age further enabled us to account for similar confounding due to higher rates of mental illness in women in certain periods in life, and the higher rates of neonatal adversity among offspring of very young / older women^14^. While healthcare in Israel is provided irrespective of demographic factors and socioeconomic status, accounting for family SES allowed us to account for potential differences due to *e*.*g*. healthcare seeking behaviors across women in SES strata. Family SES was defined as number of electronic devices per person and per capita income. Lastly, our earlier work has shown that women with mental health diagnosis in pregnancy are more likely to also receive many other physical health diagnoses^9^, and some of these diagnoses could be independently associated with neonatal adversity^10,15^. Adjusting for the burden of physical health diagnoses – defined as the total number of distinct ICD-9 level-3 diagnoses recorded during the exposure period – has allowed us to observe whether the associations between maternal psychiatric diagnosis and neonatal outcomes occur independently of these physical factors. Information about all covariates was obtained from the Meuhedet HMO records.

### Statistical Analysis

To estimate the effects of maternal mental health disorder on each neonatal outcome, we constructed a series of analogous logistic regression models iteratively controlling for covariates. in all models we adjusted for child’s year of birth and maternal age (model 1); next, we adjusted for family SES (model 2); finally, we also adjusted for the number of distinct medical comorbidities during the exposure period as a proxy measure for the maternal burden of physical comorbidities (model 3).

To investigate familial confounding, we estimated the effects of paternal mental health disorder on offspring using logistic regression. To mitigate against potential inflation of paternal results due to assortative mating^16^, we included the effects of both parental diagnoses in the model, adjusted for child’s year of birth, parental ages at child’s birth and family SES.

To account for familial correlations due to the presence of siblings in the dataset, all models were stratified by maternal ID to obtain robust standard errors. Since children from larger families were more likely to be included in the dataset (if one child is randomly selected, their siblings are also included), in all models we also adjusted for this sampling strategy by using inverse probability weights determined by the number of children born within the cohort years in the family (see^17^ for details on weights calculation).

In the secondary analyses, (a) we tested the effects of specific mental health diagnosis (specific ICD-9, 3-digit code) on birth complications and congenital anomalies. The diagnoses of mental health disorders tested in these analyses are listed in the Exposures section. Separately, (b) in order to test that our results are not due to differential physical health screening based on psychiatric disorder status, we also tested the association between maternal burden of physical health diagnoses and neonatal adversity in subsets of women with and without a diagnosis of mental health disorder. Finally, (c) to interrogate the association between maternal psychiatric diagnosis in pregnancy and more severe neonatal adversity, we repeated the analyses re-defining the outcome measures to only include the codes present in <1.5% of the sample. All statistical procedures were analogous to those performed in the main analysis, *i*.*e*. we used logistic regression with weights, calculated robust standard errors, and iteratively added the covariates.

## Results

Our primary sample consisted of 95,978 children - including both the random sample of the birth cohort and their siblings. After removing individuals based on the exclusion criteria above, our analytical sample consisted of 74,533 individuals, including 36,534 females (49% of the cohort), and 37,999 males (51% of the sample). The sample demographics are presented in **Table 1**. Among these individuals, 6,674 (9.1%) experienced complications at birth, and 14,569 (19.9%) were born with any type of a congenital anomaly. Majority of the birth complications related to perinatal jaundice (ICD-9: 774, 5.3%), umbilical cord problems (ICD-9: 663; 1.4% sample) and congenital infection (ICD-9: 771; 1.4%), while the most common congenital anomalies were musculoskeletal deformities (ICD-9:754 ; 8.2%) and other congenital anomalies of limbs (ICD-9: 755; 5.3%). After excluding the codes with a prevalence >1.5% in the sample, birth complications and congenital anomalies were diagnosed in 4.1% and 6.8% of the sample, respectively. Prevalence of each ICD-9 diagnosis included in the analyses of birth complications and congenital anomalies is presented in Supplementary Table 1.

**Table 1.** Sample Demographics.

|  | Maternal mental health diagnosis around pregnancy period |  | Paternal mental health diagnosis around pregnancy period |  |
| --- | --- | --- | --- | --- |
|  | Yes (N=3,284) | No (N=72,182) | Yes (N=4,234) | No (N=71,232) |
| % Female | 49.95% | 48.97% | 47.90% | 49.08% |
| Mean maternal age at child's birth (SD) | 31.4 (5.3) | 29.7 (5.4) | 31.0 (5.4) | 29.7 (5.4) |
| Mean paternal age at child's birth (SD) | 34.1 (6.0) | 32.3 (6.1) | 33.8 (6.1) | 32.3 (6.1) |
| Mean number of maternal diagnoses in pregnancy (SD) | 20.3 (17.5) | 10.7 (10.1) | 14.6 (13.4) | 10.9 (10.5) |
| Mean number of paternal diagnoses in pregnancy (SD) | 10.4 (12.8) | 7.4 (9.5) | 15.5 (17.5) | 7.1 (8.8) |
| Median SES (MAD) | 7.0 (4.4) | 7.0 (4.4) | 7.0 (4.4) | 7.0 (4.4) |
| N (%) offspring birth complications | 375 (11.6) | 6,299 (9.0) | 451 (10.9) | 6216 (9.0) |
| N (%) offspring congenital anomalies | 776 (24.1) | 13,793 (19.7) | 926 (22.4) | 13,629 (19.7) |

### Maternal mental health diagnosis in pregnancy period is associated with birth complications and congenital anomalies

Maternal diagnosis of any mental health disorder around the pregnancy period was significantly associated with birth complications and congenital anomalies, after adjustments for child’s year of birth, maternal age, and SES (*birth complications*: OR=1.3 (1.2-1.4), p<0.001; *congenital anomalies:* OR=1.2 (1.1-1.3), p<0.001; **Table 2**). When the neonatal outcomes were re-defined to only include the relatively rare (<1.5%) ICD-9 codes, the odds associated with maternal mental health diagnosis remained similar (*birth complications*: OR=1.4 (1.2-1.6), p<0.001; *congenital anomalies:* OR=1.4 (1.2-1.5), p<0.001; Supplementary Table S4).

**Table 2.** Association between maternal diagnosis of psychiatric disorder and (i) birth complications (upper panel); and (ii) congenital anomalies. Iterative addition of covariates in models 1-3 is described in a-c.

|  | Model 1<sup>a</sup> |  |  | Model 2<sup>b</sup> |  |  | Model 3<sup>c</sup> |  |  |
| --- | --- | --- | --- | --- | --- | --- | --- | --- | --- |
|  | OR | 95% CI | *p* | OR | 95% CI | *p* | OR | 95% CI | *p* |
| **Birth Complications** | 1.3 | 1.1-1.4 | <0.001 | 1.3 | 1.2-1.4 | <0.001 | 1.1 | 0.9-1.2 | 0.40 |
| **Congenital Anomalies** | 1.2 | 1.1-1.3 | <0.001 | 1.2 | 1.1-1.3 | <0.001 | 1.1 | 1.0-1.2 | 0.14 |

<sup>a</sup> Adjusted for maternal age and child's year of birth
<sup>b</sup> Adjusted for maternal age, child's year of birth, family SES
<sup>c</sup> Adjusted for maternal age, child's year of birth, family SES, burden of maternal physical health diagnoses

In the secondary analyses, where we analyzed the effects of specific maternal mental health disorders on neonatal adversity, anxiety disorder was associated with birth complications and congenital anomalies, while personality disorder was associated with birth complications only (anxiety - *birth complications*: OR=1.6 (1.4-1.7), p<0.001; *anomalies:* OR=1.3 (1.1-1.4), p<0.001; personality disorder – *birth complications*: OR=2.0 (1.4-2.7), p = 0.04; Supplemental Table S5). There were no statistically significant effects of the diagnosis of other mental health disorders with specific symptoms or depressive disorders on either measure of neonatal adversity (Supplemental Table S5).

### Evidence for familial confounding in the association between maternal mental health diagnosis and neonatal adversity

In a joint model testing concurrently the effects of paternal and maternal diagnosis, both were significantly associated with the study outcomes (*birth complications*: OR_mat_=1.3 (1.1-1.4), p < 0.001; OR_pat_=1.2 (1.1-1.3), p=0.004; *congenital anomalies*: OR_mat_=1.2 (1.1-1.3), p<0.001; OR_pat_=1.1 (1.0-1.2), p=0.01; **Table 3**). This pattern of results was similar in the secondary analyses of specific codes for maternal and paternal diagnoses (Supplementary Table S6). However, while the confidence intervals overlapped, the risk estimates associated with maternal psychiatric diagnosis were consistently higher compared to those associated with paternal diagnosis, both in the main and secondary analyses.

**Table 3.** Association between maternal and paternal diagnoses of psychiatric disorder and (i) birth complications (upper panel); and (ii) congenital anomalies, adjusted for child’s year of birth, maternal and paternal ages, family SES.

|  |  | OR | 95% CI | p |
| --- | --- | --- | --- | --- |
| **Birth Complications** | **maternal psych dx** | 1.3 | 1.1-1.4 | <0.001 |
|  | **paternal psych dx** | 1.2 | 1.1-1.3 | 0.004 |
| **Congenital Anomalies** | **maternal psych dx** | 1.2 | 1.1-1.3 | <0.001 |
|  | **paternal psych dx** | 1.1 | 1.0-1.2 | 0.01 |

When the outcomes were defined only based on the ICD-9 codes diagnosed in <1.5% of the sample, we observed significant effects only of maternal, but not paternal psychiatric diagnosis (*birth complications*: OR_mat_= 1.4 (1.2-1.6), p<0.001; OR_pat_= 1.1 (1.0-1.3), p=0.13; *congenital anomalies*: OR_mat_= 1.3 (1.2-1.5), p<0.001; OR_pat_= 1.1 (0.9-1.2), p=0.35; Supplementary Table S7).

### Adjustment for burden of physical comorbidities renders the effects of maternal mental health diagnosis on offspring non-significant

The associations between maternal mental health and measures of neonatal adversity – which remained significant through the adjustment for child’s year of birth, maternal age at child’s birth and family SES – were no longer statistically significant after adjusting for the burden of physical health comorbidities (model 3; **Table 2**). Meanwhile, higher number of maternal physical health diagnoses was strongly associated with the risk of neonatal adversity (*birth complications*: OR=1.2 (1.1-1.3), p<0.001; *congenital anomalies*: OR=1.1 (1.0-1.2); p=0.02). The association between the number of physical health diagnoses and neonatal adversity was present both in women with and without mental health diagnosis, suggesting that the effect was not driven only by increased health surveillance in women with psychiatric disorders (*birth complications*: OR_psych-YES_= 1.01 (1.00-1.02), p=0.04; OR_psych-NO_= 1.02 (1.02-1.02), p<0.001; *congenital anomalies*: OR_psych-YES_=1.01 (1.00-1.01), p=0.01; OR_psych-NO_=1.02 (1.01-1.02), p<0.001).

When considering only the relatively rare types of birth complications and congenital anomalies, we observed that adjusting for maternal physical comorbidities results in attenuation of the effect sizes and significance levels of both outcomes (*birth complications*: OR=1.2 (1.0-1.4), p=0.06; *congenital anomalies*: OR=1.2 (1.0-1.3); p=0.04; Supplemental Table S4), similarly to the results in the main analyses. This pattern of results was similar for nearly all specific diagnoses, with only maternal anxiety disorder remaining significantly associated with birth complications after adjustment for maternal burden of physical health diagnoses (albeit with substantially lower point estimates compared to prior to the adjustment; Supplemental Table S6).

## Discussion

Our findings replicate results from the earlier studies demonstrating that maternal diagnosis of a mental health disorder in pregnancy is associated with birth complications and congenital anomalies. We further show that the key factors contributing to neonatal adversity in offspring of women with a psychiatric diagnosis include (i) physical factors associated with maternal mental health diagnosis, and (ii) familial factors associated with both maternal and paternal mental health diagnoses. Taken together, our results underscore complex relationships between parental mental and physical health, as well as the familial factors in influencing the risk of neonatal adversity.

First, we observed that the risk of neonatal adversity associated with a maternal diagnosis of mental health disorder is attenuated (but not completely eliminated) after adjustment for the burden of physical health diagnoses. These results are consistent with our^9^ and others’^8^ earlier results demonstrating high comorbidity between mental and physical health, including during pregnancy – with likely knock-on effects on offspring outcomes. Previously, other studies have suggested the role of increased inflammation^5,6,18–20^ or lifestyle factors, *e*.*g*. smoking^21^ as potential mediators of the observational association between maternal mental health and neonatal outcomes. Our findings demonstrating the importance of physical health are consistent with these suggestions: factors like maternal inflammation or smoking, and the burden of physical health conditions in our study, could be on the same causal pathway between maternal mental health and neonatal outcomes. While in the absence of more detailed phenotypic data we could not test this directly, physiological correlates of physical comorbidities of psychiatric disorders – including *e*.*g*. immune, metabolic and hormonal factors - should be further interrogated in the future.

While the earlier studies typically suggested a *mediating* effect of inflammation and lifestyle – whereby mental health disorder leads to increased inflammation and/or lifestyle changes, which then in turn lead to offspring adversity^20^ – we currently lack the scope to distinguish such putative mediating, from confounding effects (i.e. whereby mental health is the consequence, not a cause of *e*.*g*. increased inflammation). In the future, determining the up- and downstream nature of the associations between mental and physical health will be critical for identification of the potentially modifiable factors influencing risk of neonatal adversity. Nevertheless, irrespective of the direction of such causal effects, our findings suggest that stronger emphasis on physical health is warranted in both the studies exploring the effects of maternal mental health on offspring, and in clinical settings.

Secondly, as paternal exposures cannot directly influence the fetus – creating a so-called negative control - performing analogous analyses in relation to paternal diagnosis of a mental health disorder allowed us to highlight the potential familial confounding underlying the observed associations. Such familial factors can include both genetic and environmental correlates of mental illness, including *e*.*g*. parental socioeconomic factors^22^, urbanicity^23^ or pollution^24^ in the area – each of which has also been linked with offspring adversity^21^. Similarly as with regard to maternal physical health, future studies incorporating a wider range of measures of the home environment and genetic factors should further examine the exact biological processes through which these factors could act to affect the child. Importantly, our strategy of estimating the risk of both maternal and paternal exposures jointly mitigates against potential confounding due to assortative mating^16^, i.e. whereby propensity for mental health disorder in a parent increases the probability of choosing a partner who is also more likely to suffer from it. In secondary analyses we observed that the role of familial confounding was lower for the relatively rare (diagnosed in <1.5% children in our sample) birth complications and neonatal anomalies. This raises the possibility that these – presumably more severe – outcomes are to a lesser extent influenced by familial factors.

Crucially, neither maternal physical health nor familial factors linked with mental health diagnosis appeared to on their own fully explain the observed effects, suggesting their joint effects on the risk of neonatal adversity. We infer this from (i) the point estimates associated with the effects of maternal diagnosis being attenuated, but still indicated increased risk, following the adjustment for physical health; and (ii) the estimates of risk for neonatal adversity associated with maternal diagnosis being consistently higher than those associated with paternal diagnosis.

The strengths of this study include use of a large, population-based sample, and reliance on the data from one of a largest Israeli HMO, limiting bias due to ascertainment or differential access to healthcare across individuals in our sample. Using the registry-based medical enabled us to ascertain both exposure and outcome with a high degree of confidence – mitigating against the problems associated with *e*.*g*. retrospective recall of mental health during pregnancy by the parents, or missing the neonatal outcomes that might have not been clinically visible immediately at birth. Availability of rich paternal health data allowed us to interrogate familial confounding, and account for the potential inflation of the results due to assortative mating. Finally, inclusion of a wide range of covariates available through the HMO records, and accounting for the sampling strategy, represent additional strengths of our work. However, we also acknowledge the limitations, including lack of a replication sample, availability of only a compound measure of family SES and limited medical data, precluding testing the potential physiological underpinnings of the observed effects.

Future research should further investigate the specific parental factors that in our analyses were captured by maternal burden of comorbidities - including specific diagnoses that contribute to these associations, and the finer underlying mechanisms. In the meantime, studies exploring the potential effects of maternal mental health problems in pregnancy should account for the presence of comorbid physical health diagnoses - which, as we have previously shown^9^ are considerably more common in pregnant women with a mental health diagnosis.

In conclusion, our results indicate that maternal diagnosis of mental health disorder around pregnancy is associated with an increased risk of neonatal adversity, including birth complications and neonatal anomalies. These associations reflect contributions of the maternal physical health, and familial factors, associated with a diagnosis of mental health disorder.

## Data Availability

Data access rules do not permit public sharing of the data. Interested researchers should discuss access options with Arad Kodesh and Stephen Levine.

## Supplemental Tables

**Table S1.** Prevalence of specific ICD-9 diagnoses of birth complications and congenital anomalies in the sample.

| Birth complications |  |
| --- | --- |
| Diagnosis (ICD-9) | Prevalence<br>n (%) |
| Obstructed labor (660) | 0 (0) |
| Abnormality of forces of labor (661) | 27 (<0.1) |
| Long labor (662) | 0 (0) |
| Umbilical cord complications during labor and delivery (663) | 1005 (1.4) |
| Trauma to perineum and vulva during delivery (664) | 9 (<0.1) |
| Other obstetrical trauma (665) | 3 (<0.1) |
| Postpartum hemorrhage (666) | 0 (0) |
| Retained placenta or membranes without hemorrhage (667) | 0 (0) |
| Complications of the administration of anesthetic or other sedation in labor and delivery (668) | 0 (0) |
| Other complications of labor and delivery not elsewhere classified (669) | 113 (0.2) |
| Other fetal conditions (678) | 0 (0) |
| Complications of in utero procedures (679) | 0 (0) |
| Fetus or newborn affected by maternal conditions which may be unrelated to present pregnancy (760) | 0 (0) |
| Fetus or newborn affected by maternal complications of pregnancy (761) | 0 (0) |
| Fetus or newborn affected by complications of placenta cord and membranes (762) | 0 (0) |
| Fetus or newborn affected by other complications of labor and delivery (763) | 0 (0) |
| Slow fetal growth and fetal malnutrition (764) | 81 (0.1) |
| Disorders relating to short gestation and low birthweight (765) | 553 (0.7) |
| Disorders relating to long gestation and high birthweight (766) | 35 (<0.1) |
| Birth trauma (767) | 144 (0.2) |
| Intrauterine hypoxia and birth asphyxia (768) | 13 (<0.1) |
| Respiratory distress syndrome in newborn (769) | 71 (0.1) |
| Other respiratory conditions of fetus and newborn (770) | 78 (0.1) |
| Infections specific to the perinatal period (771) | 592 (0.8) |
| Fetal and neonatal hemorrhage (772) | 100 (0.1) |
| Hemolytic disease of fetus or newborn due to isoimmunization (773) | 18 (<0.1) |
| Other perinatal jaundice (774) | 3917 (5.3) |
| Endocrine and metabolic disturbances specific to the fetus and newborn (775) | 47 (<0.1) |
| Hematological disorders of newborn (776) | 37 (0.1) |
| Perinatal disorders of digestive system (777) | 18 (<0.1) |
| Conditions involving the integument and temperature regulation of fetus and newborn (778) | 240 (0.3) |
| Other and ill-defined conditions originating in the perinatal period (779) | 202 (0.3) |
| Congenital anomalies |  |
| Diagnosis (ICD-9) | Prevalence<br>n (%) |
| Anencephalus and similar anomalies (740) | 1 (<0.1) |
| Spina bifida (741) | 41 (0.1) |
| Other congenital anomalies of nervous system (742) | 298 (0.4) |
| Congenital anomalies of eye (743) | 276 (0.4) |
| Congenital anomalies of ear face and neck (744) | 182 (0.2) |
| Bulbus cordis anomalies and anomalies of cardiac septal closure (745) | 853 (1.2) |
| Other congenital anomalies of heart (746) | 183 (0.2) |
| Other congenital anomalies of circulatory system (747) | 308 (0.4) |
| Congenital anomalies of respiratory system (748) | 531 (0.7) |
| Cleft palate and cleft lip (749) | 77 (0.1) |
| Other congenital anomalies of upper alimentary tract (750) | 996 (1.4) |
| Other congenital anomalies of digestive system (751) | 66 (0.1) |
| Congenital anomalies of genital organs (752) | 1627 (2.2) |
| Congenital anomalies of urinary system (753) | 204 (0.3) |
| Certain congenital musculoskeletal deformities (754) | 6036 (8.2) |
| Other congenital anomalies of limbs (755) | 3858 (5.3) |
| Other congenital musculoskeletal anomalies (756) | 561 (0.8) |
| Congenital anomalies of the integument (757) | 750 (1.0) |
| Chromosomal anomalies (758) | 141 (0.2) |
| Other and unspecified congenital anomalies (759) | 293 (0.4) |

**Table S2.** Prevalence of birth complications and congenital anomalies in relation to maternal diagnosis of mental health disorder.

|  |  | Birth complications, n (%) | Congenital anomalies, n (%) |
| --- | --- | --- | --- |
| anxiety, dissociative, and somatoform disorders (ICD-9: 300) | yes | 203 (13.4) | 375 (24.8) |
|  | no | 6,471 (9) | 14,194 (19.8) |
| personality disorders (ICD-9: 301) | yes | 13 (17.1) | 23 (30.3) |
|  | no | 6,661 (9.1) | 14,546 (19.9) |
| other mental health disorders with specific symptoms (ICD-9: 307) | yes | 72 (7.9) | 211 (23.2) |
|  | no | 6,602 (9.1) | 14,358 (19.8) |
| depressive disorders (ICD-9: 311) | yes | 65 (10.7) | 145 (23.8) |
|  | no | 6,609 (9.1) | 14,424 (19.9) |
| Any mental health diagnosis |  | 375 (11.6) | 776 (24.1) |
| No mental health diagnosis |  | 6,299 (9.0) | 13,793 (19.7) |

**Table S3.**
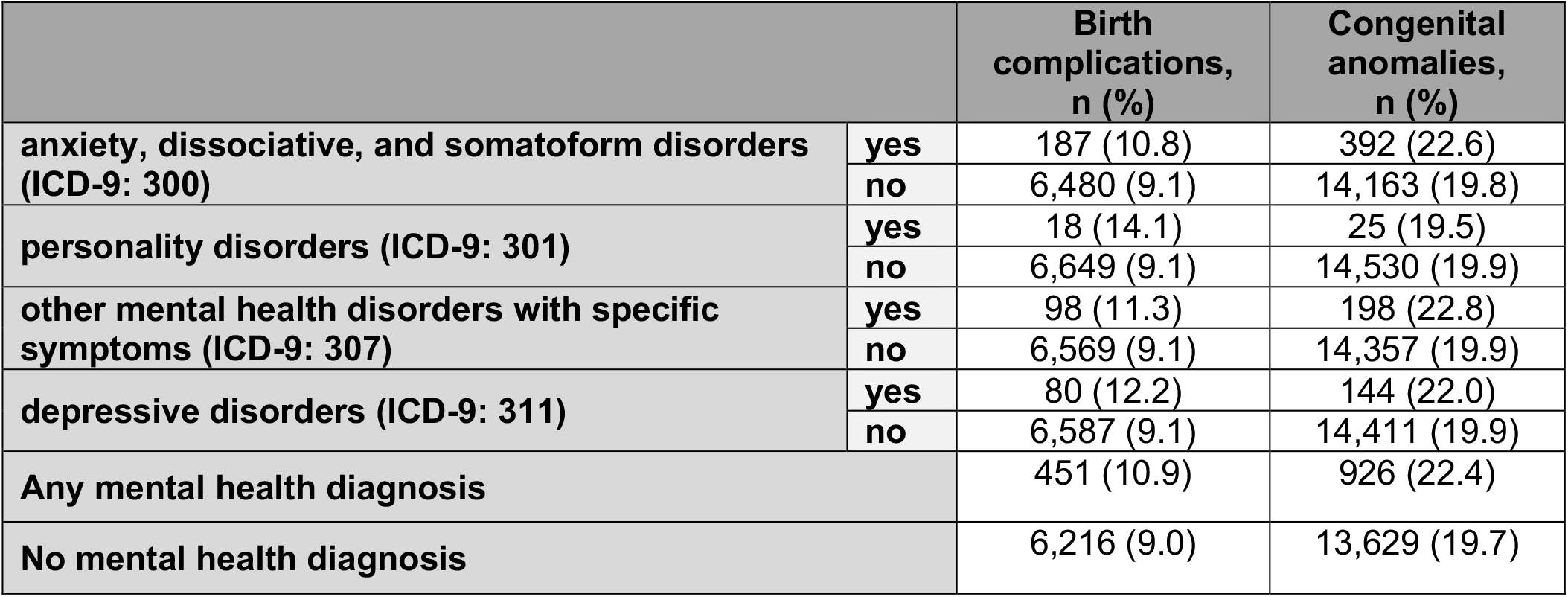
Prevalence of birth complications and congenital anomalies in relation to paternal diagnosis of mental health disorder.

|  |  | Birth complications, n (%) | Congenital anomalies, n (%) |
| --- | --- | --- | --- |
| anxiety, dissociative, and somatoform disorders (ICD-9: 300) | yes | 187 (10.8) | 392 (22.6) |
|  | no | 6,480 (9.1) | 14,163 (19.8) |
| personality disorders (ICD-9: 301) | yes | 18 (14.1) | 25 (19.5) |
|  | no | 6,649 (9.1) | 14,530 (19.9) |
| other mental health disorders with specific symptoms (ICD-9: 307) | yes | 98 (11.3) | 198 (22.8) |
|  | no | 6,569 (9.1) | 14,357 (19.9) |
| depressive disorders (ICD-9: 311) | yes | 80 (12.2) | 144 (22.0) |
|  | no | 6,587 (9.1) | 14,411 (19.9) |
| Any mental health diagnosis |  | 451 (10.9) | 926 (22.4) |
| No mental health diagnosis |  | 6,216 (9.0) | 13,629 (19.7) |

**Table S4.**
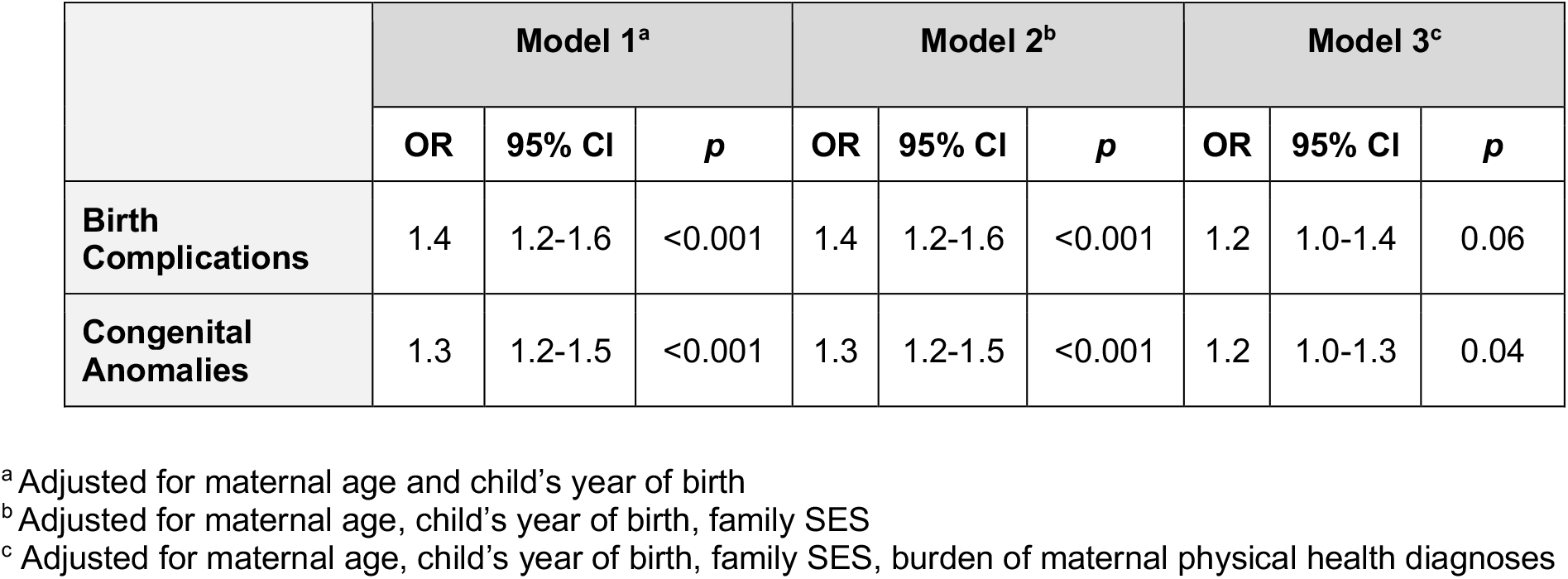
Association between maternal diagnosis of psychiatric disorder and (i) birth complications (upper panel); and (ii) congenital anomalies, after exclusion of the ICD-9 codes diagnosed in >1.5% of the sample. Iterative addition of covariates in models 1-3 is described in a-c.

|  | Model 1<sup>a</sup> |  |  | Model 2<sup>b</sup> |  |  | Model 3<sup>c</sup> |  |  |
| --- | --- | --- | --- | --- | --- | --- | --- | --- | --- |
|  | OR | 95% CI | *p* | OR | 95% CI | *p* | OR | 95% CI | *p* |
| Birth Complications | 1.4 | 1.2-1.6 | <0.001 | 1.4 | 1.2-1.6 | <0.001 | 1.2 | 1.0-1.4 | 0.06 |
| Congenital Anomalies | 1.3 | 1.2-1.5 | <0.001 | 1.3 | 1.2-1.5 | <0.001 | 1.2 | 1.0-1.3 | 0.04 |

<sup>a</sup> Adjusted for maternal age and child's year of birth
<sup>b</sup> Adjusted for maternal age, child's year of birth, family SES
<sup>c</sup> Adjusted for maternal age, child's year of birth, family SES, burden of maternal physical health diagnoses

**Table S5.**
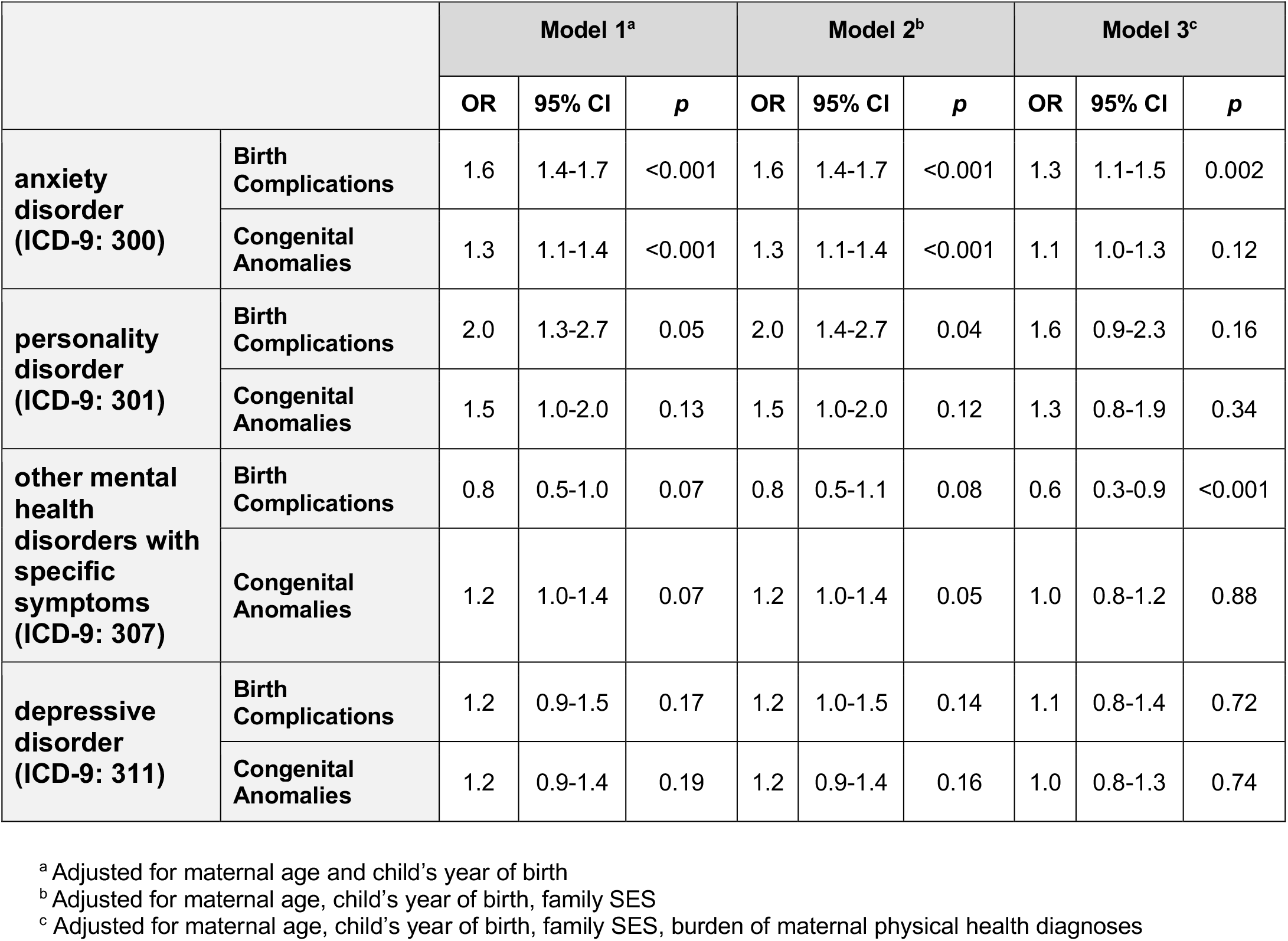
Association between maternal diagnosis of anxiety disorder (ICD-9: 300), personality disorder (ICD-9: 301), other mental health disorders with specific symptoms (ICD-9: 307) and depressive disorder (ICD-9: 311) with (i) birth complications (upper panel); and (ii) congenital anomalies. Iterative addition of covariates in models 1-3 is described in a-c.

|  |  | Model 1<sup>a</sup> |  |  | Model 2<sup>b</sup> |  |  | Model 3<sup>c</sup> |  |  |
| --- | --- | --- | --- | --- | --- | --- | --- | --- | --- | --- |
|  |  | OR | 95% CI | *p* | OR | 95% CI | *p* | OR | 95% CI | *p* |
| **anxiety disorder (ICD-9: 300)** | **Birth Complications** | 1.6 | 1.4-1.7 | <0.001 | 1.6 | 1.4-1.7 | <0.001 | 1.3 | 1.1-1.5 | 0.002 |
|  | **Congenital Anomalies** | 1.3 | 1.1-1.4 | <0.001 | 1.3 | 1.1-1.4 | <0.001 | 1.1 | 1.0-1.3 | 0.12 |
| **personality disorder (ICD-9: 301)** | **Birth Complications** | 2.0 | 1.3-2.7 | 0.05 | 2.0 | 1.4-2.7 | 0.04 | 1.6 | 0.9-2.3 | 0.16 |
|  | **Congenital Anomalies** | 1.5 | 1.0-2.0 | 0.13 | 1.5 | 1.0-2.0 | 0.12 | 1.3 | 0.8-1.9 | 0.34 |
| **other mental health disorders with specific symptoms (ICD-9: 307)** | **Birth Complications** | 0.8 | 0.5-1.0 | 0.07 | 0.8 | 0.5-1.1 | 0.08 | 0.6 | 0.3-0.9 | <0.001 |
|  | **Congenital Anomalies** | 1.2 | 1.0-1.4 | 0.07 | 1.2 | 1.0-1.4 | 0.05 | 1.0 | 0.8-1.2 | 0.88 |
| **depressive disorder (ICD-9: 311)** | **Birth Complications** | 1.2 | 0.9-1.5 | 0.17 | 1.2 | 1.0-1.5 | 0.14 | 1.1 | 0.8-1.4 | 0.72 |
|  | **Congenital Anomalies** | 1.2 | 0.9-1.4 | 0.19 | 1.2 | 0.9-1.4 | 0.16 | 1.0 | 0.8-1.3 | 0.74 |

<sup>a</sup> Adjusted for maternal age and child's year of birth
<sup>b</sup> Adjusted for maternal age, child's year of birth, family SES
<sup>c</sup> Adjusted for maternal age, child's year of birth, family SES, burden of maternal physical health diagnoses

**Table S6.**
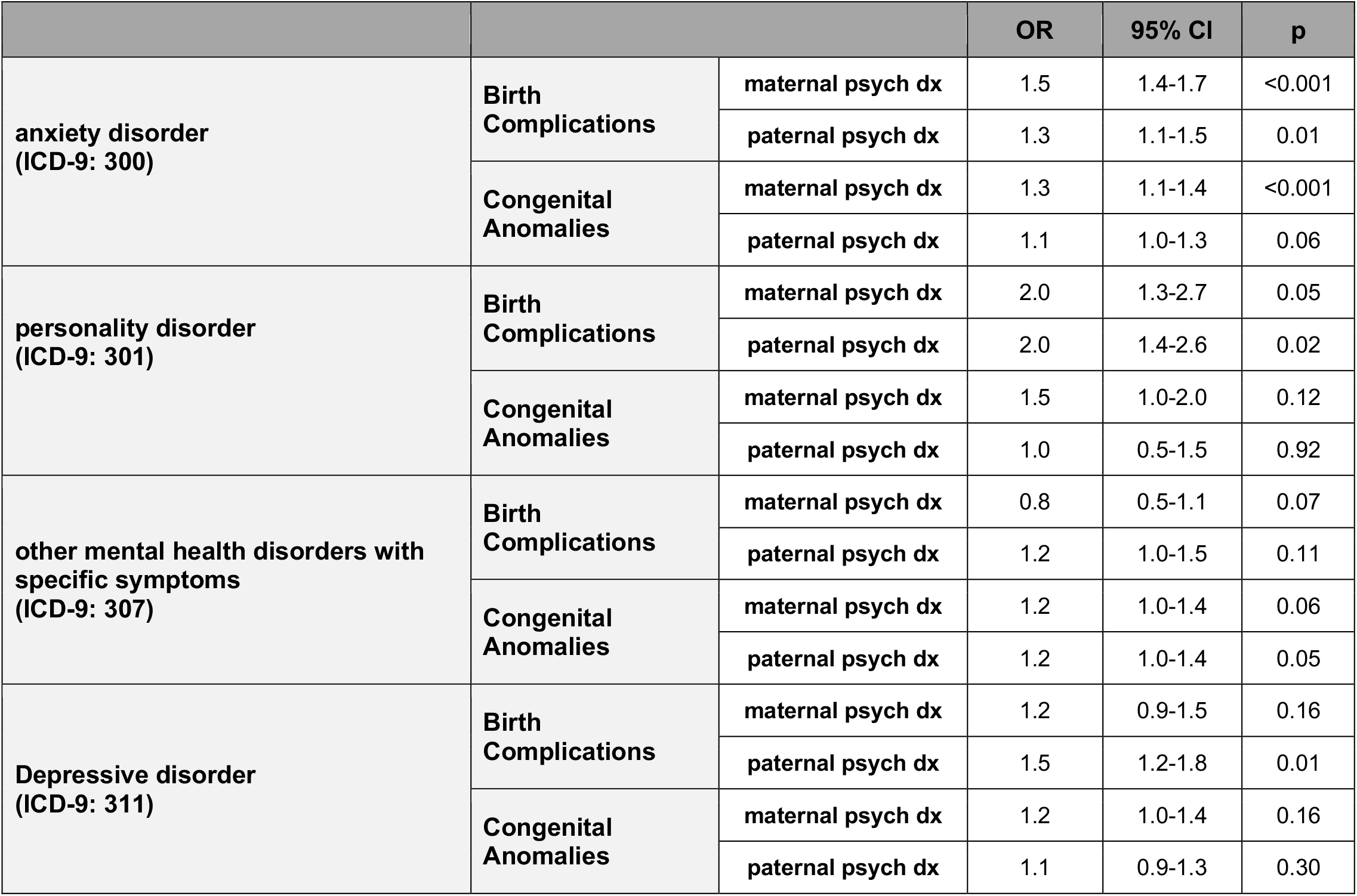
Association between maternal and paternal diagnosis of anxiety disorder (ICD-9: 300), personality disorder (ICD-9: 301), other mental health disorders with specific symptoms (ICD-9: 307) and depressive disorder (ICD-9: 311) with birth complications and congenital anomalies; within exposure-outcome pair, maternal and paternal effects were estimated jointly, adjusting for child’s year of birth, maternal and paternal ages, family SES.

**Table S7.** Association between maternal and paternal diagnoses of psychiatric disorder and (i) birth complications (upper panel); and (ii) congenital anomalies, after exclusion of the ICD-9 codes diagnosed in >1.5% of the sample. All analyses were adjusted for child’s year of birth, maternal and paternal ages, family SES.

|  |  | OR | 95% CI | p |
| --- | --- | --- | --- | --- |
| Birth<br>Complications | maternal psych dx | 1.4 | 1.2-1.6 | <0.001 |
|  | paternal psych dx | 1.1 | 1.0-1.3 | 0.13 |
| Congenital<br>Anomalies | maternal psych dx | 1.3 | 1.2-1.5 | <0.001 |
|  | paternal psych dx | 1.1 | 0.9-1.2 | 0.35 |

